# Considerations for ensuring safety during telerehabilitation of people with stroke. A protocol for a scoping review

**DOI:** 10.1101/2022.10.06.22280769

**Authors:** Ruvistay Gutierrez-Arias, Camila González-Mondaca, Vinka Marinkovic-Riffo, Marietta Ortiz-Puebla, Fernanda Paillán-Reyes, Pamela Seron

**Affiliations:** Servicio de Medicina Física y Rehabilitación, Unidad de Kinesiología, Instituto Nacional del Tórax, Santiago, Chile; Exercise and Rehabilitation Sciences Institute, School of Physical Therapy, Faculty of Rehabilitation Sciences, Universidad Andres Bello, Santiago, 7591538, Chile; Centro de Excelencia CIGES, Universidad de La Frontera, Temuco, Chile; Departamento de Ciencias de la Rehabilitación, Facultad de Medicina, Universidad de La Frontera, Temuco, Chile

**Author notes:** Corresponding author: Ruvistay Gutierrez-Arias, PT MSc. Servicio de Medicina Física y Rehabilitación, Unidad de Kinesiología, Instituto Nacional del Tórax, Santiago, Chile & Exercise and Rehabilitation Sciences Institute, School of Physical Therapy, Faculty of Rehabilitation Sciences, Universidad Andres Bello, Santiago, 7591538, Chile. Address all correspondence to.

## Abstract

**Introduction:** Exercise interventions have a positive impact on people with stroke. However, access to exercise interventions is variable, and there may be a delay in the start of rehabilitation. Telerehabilitation has enabled the delivery of exercise interventions replacing the traditional face-to-face approach. Aspects related to the safety of people with stroke should be considered to avoid adverse events during the delivery of exercise interventions remotely. However, such information is scattered in the literature, and the detail with which measures taken during the implementation of exercise interventions for people with stroke are reported is unknown.

**Objective:** To summarise measures or aspects targeted at reducing the incidence of adverse events during the delivery of exercise interventions through telerehabilitation in patients after stroke.

**Materials and methods:** A scoping review will be conducted. A systematic search in MEDLINE-Ovid, Embase-Ovid CENTRAL, CINAHL Complete (EBSCOhost), and other resources will be carried out. We will include primary studies, published in full text in any language, involving people with stroke who undergo telerehabilitation where exercise is the main component. Two reviewers will independently select studies and extract data, and disagreements will be resolved by consensus or a third reviewer. The results will be reported in a narrative form, using tables and figures to support them.

**Discussion:** To implement this strategy within rehabilitation services, one of the first aspects to be solved is to ensure the safety of people. The results of this scoping review could contribute an information base for clinicians and decision-makers when designing remotely delivered exercise intervention programs.

**Registration number:** INPLASY202290104.

## Introduction

Stroke is a disease characterized by a focal deficit due to acute injury to the central nervous system (1). This injury of vascular origin includes cerebral infarction and intracerebral or subarachnoid hemorrhage (1). Stroke is the second leading cause of death and disability worldwide, resulting in a high burden of disease, especially in low- and middle-income countries (2). The reported prevalence of stroke globally in 2016 was 80.1 million (95% CI 74.1–86.3) (3).

The sequelae in people with stroke are diverse (4). Regarding physical function post-stroke, functional impairment of the upper and lower extremities is common, which may be due to weakness or paralysis, sensory loss, spasticity, and abnormal motor synergies (5). In addition, a near 15% prevalence of sarcopenia has been found in people with stroke (6). Gait impairment has been observed in a high percentage of people with stroke (7,8), a dysfunction that may persist despite rehabilitation (9).

More than 50% of people with stroke may experience limitations in activities such as shopping, housework, and difficulty reintegrating into community life within 6 months (10). These restrictions can result in a diminished health-related quality of life (10–12).

Rehabilitation is a multiple and comprehensive intervention, of which exercise is one of the main components. Exercise interventions have a positive impact on people with stroke (13), with a small to moderate effect on the quality of life (14), and an increase in cognitive performance (15). However, access to exercise interventions is variable (16–18), and there may be a delay in the start of physical rehabilitation (19). This could be made worse in contexts such as the COVID-19 pandemic, where health services have had to adapt and implement strategies that allow for continued service delivery, such as telerehabilitation (20).

Telerehabilitation has enabled the delivery of exercise interventions replacing the traditional face-to-face approach in patient-rehabilitator interaction (21). The potential for telerehabilitation to achieve similar clinical outcomes to traditional rehabilitation, and better than no rehabilitation at all (22), should prompt healthcare facilities to evaluate implementing remote delivery of exercise interventions for people with stroke.

For this, in addition to logistics and costs, aspects related to the safety of people with stroke should be considered to avoid adverse events during the delivery of exercise interventions. This information could be reported from studies that have evaluated the feasibility, safety, or effectiveness of telerehabilitation in this population. However, such information is scattered in the literature, and the detail with which measures taken during the implementation of exercise interventions for people with stroke are reported is unknown. Therefore, this study aims to summarise measures or aspects targeted at reducing the incidence of adverse events during the delivery of exercise interventions through telerehabilitation in patients after stroke.

## Materials and methods

A scoping review will be conducted following the updated recommendations of the Joanna Briggs Institute (JBI) (23). The protocol for this review was registered on the International Platform of Registered Systematic Review and Meta-analysis Protocols (INPLASY) under the number INPLASY202290104. The results will be reported following the Extension for Scoping Reviews of the Preferred Reporting Items for Systematic Reviews and Meta-analyses statement (PRISMA-ScR) (24).

### Search strategy

A systematic search of MEDLINE, through the Ovid platform; Embase, through the Ovid platform; Cochrane Collaboration Central Register of Controlled Trials (CENTRAL), through the Cochrane Library; and Cumulative Index of Nursing and Allied Literature Complete (CINAHL Complete), through the EBSCOhost platform. The strategy will consider a sensitive approach and the use of controlled language (MeSH, EMTREE, CINAHL Subject Heading) and natural language. The strategy to be used for MEDLINE-Ovid will be adapted to construct the search in the other databases (Table 1).

**Table 1.**
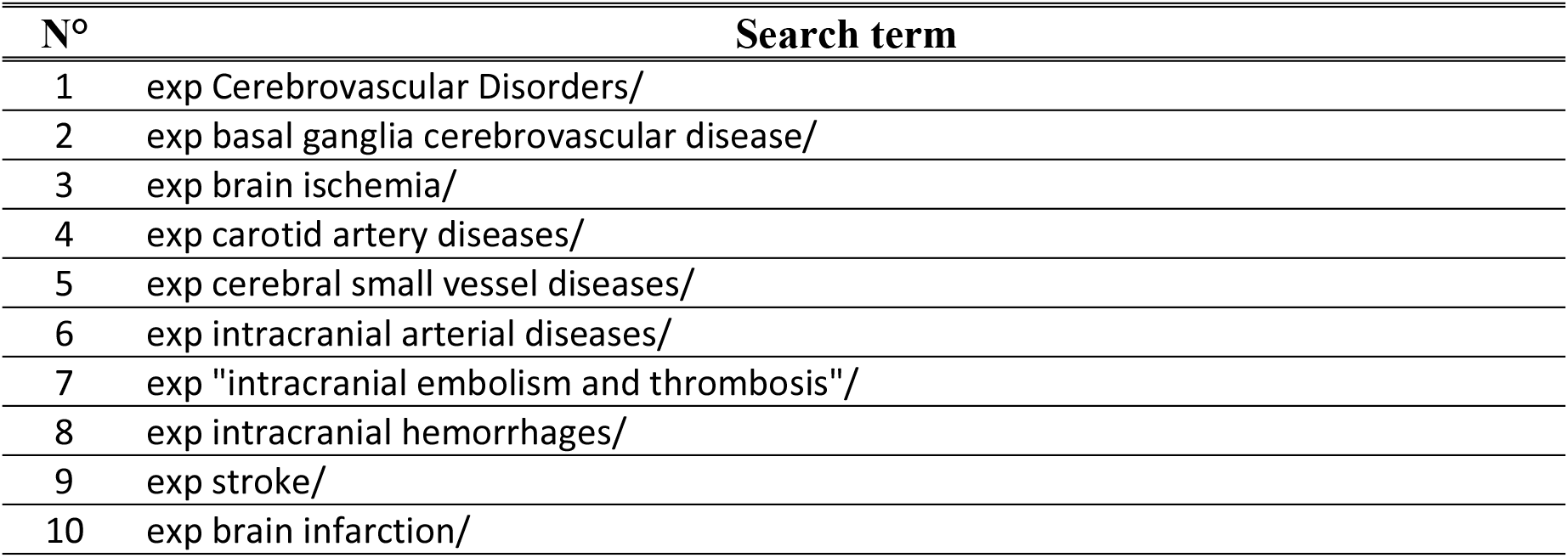
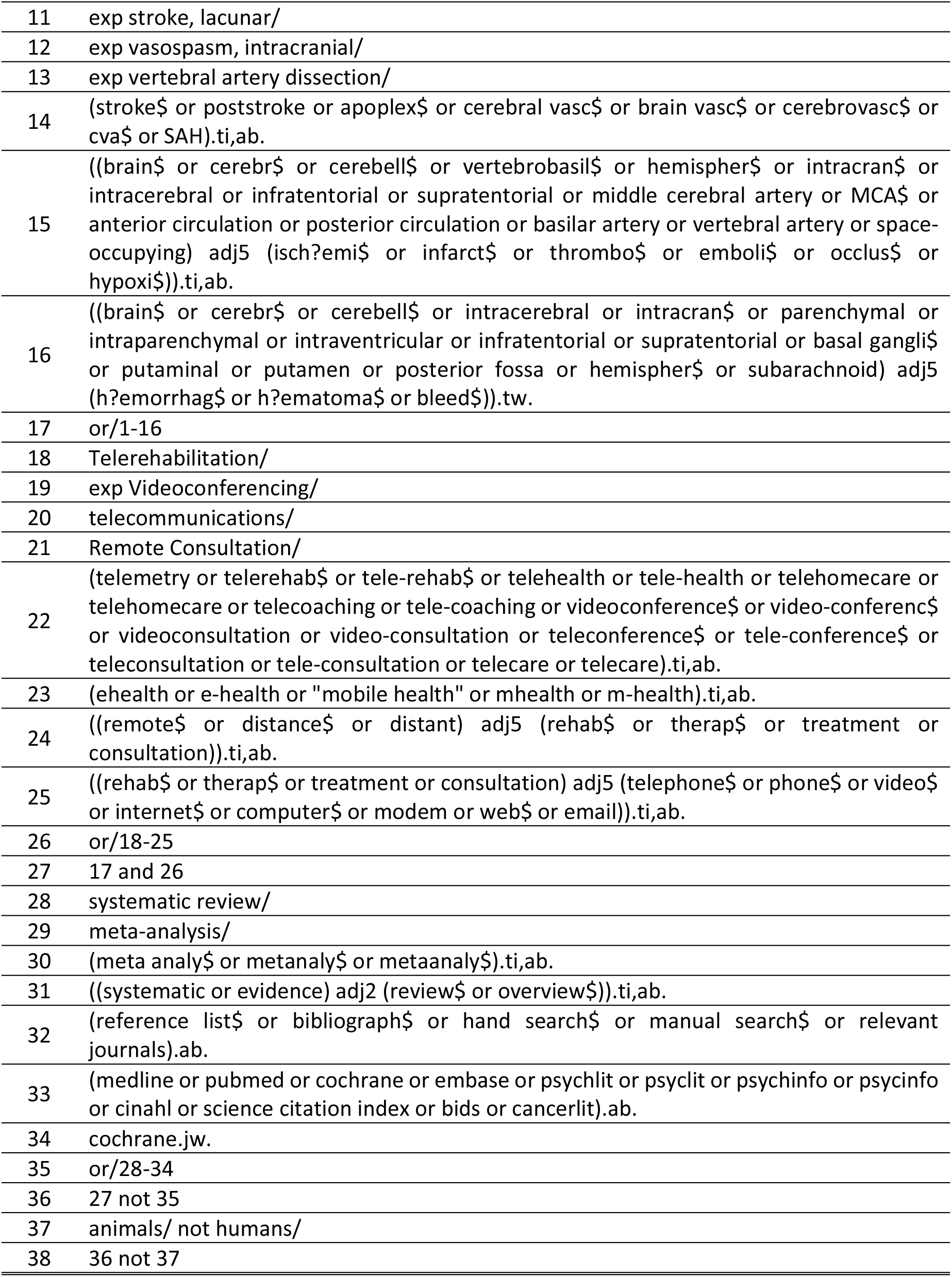
Search strategy for MEDLINE using the Ovid platform

The search was not limited by publication date, publication status, or the language of the studies. Filters will be applied to the different strategies to exclude systematic reviews, with and without meta-analysis, from the search results.

In addition, studies included in systematic reviews aimed at evaluating the effectiveness of telerehabilitation in stroke patients will be screened.

### Eligibility criteria

Eligibility criteria for study selection will be divided into participants or populations included in the studies, the concept or phenomenon involved, and the context in which the studies were conducted (PCC framework) (23). In addition, the design of the studies will be considered for inclusion in this review.

#### Participants

This review will include studies involving people with stroke, regardless of type, cause, the time course of the disease and sequelae caused.

#### Concept

This review will include studies where exercise interventions are delivered through telerehabilitation. Exercise interventions shall be understood as a subcategory of physical activity that is planned, structured, repetitive, and purposefully focused on improving or maintaining one or more components of physical fitness (25).

Interventions may be delivered synchronously, asynchronously, or mixed. Studies in which the professional delivering the intervention has face-to-face contact with the person with stroke to conduct assessments or educational sessions on the exercises to be performed (hybrid programmes) will not be excluded. The methodology for delivering interventions remotely may be by videoconferencing, mobile phone, pre-recorded videos, text message reminders, web platforms or apps.

#### Context

This review will include studies in which people with stroke perform the exercises as prescribed outside the hospital setting, either at home or in community centres, and can be performed individually or in groups. The therapist may or may not be remotely supervising the exercise sessions.

#### Study designs

Primary studies (randomised or non-randomised clinical trials, cohort studies, case-control, cross-sectional, and case reports, among others) will be included. In terms of publication status, studies in which only the abstract is available, such as those presented in conference proceedings, will be excluded. In addition, survey-based studies assessing barriers and facilitators of telerehabilitation in people with stroke will be excluded.

Studies that do not report a clinical outcome, such as level of function, quality of life, muscle strength, safety, among others, as well as the level of satisfaction, will be excluded.

The language as well as the publication date of the studies will not limit their inclusion.

### Selection of studies

Once the search for studies has been conducted, titles and abstracts will be independently screened by two research team members, who will discard studies irrelevant to this review. Subsequently, the full texts of the potential studies to be included will be analyzed to determine which articles meet all the eligibility criteria. The Rayyan® app will be used for this stage (26).

In the first instance, disagreements will be resolved by consensus, and if they persist, a third reviewer will determine the inclusion of the studies.

### Information extraction

Two reviewers will independently extract information from the included studies. An extraction form specifically designed to meet the objectives of this review will be used and will be developed in a Microsoft Excel® spreadsheet.

The information to be extracted will include aspects related to the characteristics of the publications and studies, as well as the population, interventions delivered (type of exercise and technological media used), and outcomes assessed (Table 2). In studies reporting on measures implemented to prevent adverse events during telerehabilitation sessions, detailed information will be extracted, such as the time of delivery of the intervention, the professional involved, the specific measure implemented, among others.

**Table 2.** Information to be extracted from the included studies

| Information | Description |
| --- | --- |
| Identification of the studies | Title of the study, name of the journal, year of publication, authors' names, and authors' nationality. |
| Classification of studies | Aim of the study, study design, and inclusion and exclusion criteria. |
| Exercise interventions | Type of exercise, dosage, duration of the session and the complete program, and prescribing professional. |
| Method used for telerehabilitation | Synchronicity and technology used to deliver the intervention. |
| Outcomes | List of outcomes assessed and reported in the study, other than adverse events. |
| Adverse events | Incidence and description of adverse events arising from telerehabilitation. |
| Conclusions | Conclusions related to the safety of delivering exercise interventions remotely. |

In the first instance, disagreements will be resolved by consensus, and if they persist, a third reviewer will determine the inclusion of the studies.

### Synthesis of information

The results of the search and selection of studies will be reported through a PRISMA flow chart (27). In addition, the reasons for the exclusion of full-text evaluated studies will be reported in a table.

The results will be reported in narrative form, and tables and figures will be used to synthesize the information. Waffle charts (28), or similar, will be used to represent the different primary study designs included, and the frequency of reporting of measures implemented to prevent adverse events during telerehabilitation. In addition, the frequency of the different measures implemented will be reported in one or more figures.

## Discussion

Different situations, such as the pandemic caused by COVID-19, may require the restructuring of health services to maintain people’s health care, such as rehabilitation interventions (20).

Telerehabilitation appears as an alternative that can contribute to this challenge, especially for people with stroke, due to their high disease burden (2). To implement this strategy within rehabilitation services, one of the first aspects to be solved is to ensure the safety of people (29,30).

The results of this scoping review could contribute an information base for clinicians and decision-makers when designing remotely delivered exercise intervention programs. In addition, knowledge gaps will be observed, which will serve as a basis for future research on this important topic.

## Data Availability

No datasets were generated or analysed during the current study. All relevant data from this study will be made available upon study completion.

## Acknowledgments

None

